# Evaluation of production lots of a rapid point-of-care lateral flow serological test intended for identification of IgM and IgG against the N-terminal part of the spike protein (S1) of SARS-CoV-2

**DOI:** 10.1101/2020.08.27.20182923

**Authors:** Tove Hoffman, Linda Kolstad, Bengt Rönnberg, Åke Lundkvist

## Abstract

**Background and objectives:** Several antibody tests are available to detect SARS-CoV-2 specific antibodies, many of which address different antigens. Rapid point-of-care (POC) tests have been doubted due to an eventual risk of production errors, although it is unstudied whether such error would affect test sensitivity and/or specificity. We aimed to evaluate two separate production lots of a commercially available test intended for rapid detection of IgM and IgG against the N-terminal part of the SARS-CoV-2 spike protein (S1).

**Materials and methods:** Serum samples from individuals with confirmed SARS-CoV-2 infection, by RT-PCR and/or serology, and pre-COVID-19 negative control sera gathered from a biobank during 2018 were collected. The presence of anti-S1 IgM/IgG was verified by an in-house Luminex-based serological assay, serving as reference method. The index test was a commercially available rapid POC-test (the COVID-19 IgG/IgM Rapid Test Cassette [Zhejiang Orient Gene Biotech Co Ltd, Huzhou, Zhejiang, China/Healgen Scientific, LLC, U.S.A.]).

**Results:** One hundred samples were verified positive for anti-S1 IgG (median fluorescence intensity (MFI) ≥900) and 74 for anti-S1 IgM (MFI ≥700), confirmed by RT-PCR (n=90) and/or serology (n=89). None of the negative controls (n=200; MFI <300) had SARS-CoV-2 anti-S1 IgM, while one tested positive for SARS-CoV-2 anti-S1 IgG. For the two lots, the sensitivities of the rapid test were 93.2% (69/74; 95% CI: 85.1% – 97.1%) and 87.8% (65/74; 95% CI: 78.5% – 93.5%) for IgM, respectively 93.0% (93/100; 95% CI: 86.3% – 96.6%) and 100.0% for IgG (100/100; 95% CI: 96.3% – 100.0%). The specificity for both lots was 100% for IgM (200/200; 95% CI: 98.1% – 100%) and 99.5% for IgG (199/200; 95% CI: 97.2% – 99.9%). The positive predictive value was 100% for IgM and 98.9% and 99.0% for IgG. The negative predictive value was 95.7% and 97.6% for IgM, and 96.6% and 100.0% for IgG.

**Conclusion:** The rapid POC-test used in this study is suitable to assess SARS-CoV-2 anti-S1 specific IgM/IgG, as a measure of previous virus exposure on an individual level. While the specificity was not affected by production lot, external validation of separate lots of rapid POC-tests is encouraged to ensure high sensitivity before market introduction.

## Background

In late 2019, a novel coronavirus causing severe acute respiratory disease was reported from Wuhan, Hubei Province, China. Within months, the disease COVID-19, caused by the novel severe acute respiratory syndrome coronavirus-2 (SARS-CoV-2) spread to cause a global pandemic, infecting over 23 million individuals worldwide, as of August 25.^1^ Diagnostic accuracy is vital, as the disease can resemble other viruses and bacteria and cause a wide range of symptoms; from asymptomatic infections, to those of a mild common cold or more severe symptoms such as acute respiratory distress syndrome or multi organ failure.^2, 3^ The virus is an enveloped, single-stranded RNA virus of the Coronavirus family. These viruses share structural similarities and are composed of 16 non-structural proteins and four structural proteins: the spike (S), envelope, membrane, and nucleocapsid (N) proteins. In turn, the N-terminal part of the S protein (S1) contains a receptor-binding domain (RBD) that specifically recognizes the angiotensin-converting enzyme 2 (ACE2) receptor in humans, which SARS-CoV-2 has been identified to bind to in order to infect the human host.^4-6^

There is convincing evidence that a majority of patients with past COVID-19 develop neutralizing antibodies against the virus.^7^ For detection of antibodies, there are several available serological assays developed to identify individuals with recent and past exposure and to assess the extent of exposure in a population. In turn, this might help to decide on application, enforcement or relaxation of containment measures. As proper neutralization tests are cumbersome, time-consuming and require biosafety level 3 laboratories, there are several available tests that address different SARS-CoV-2 specific antigens, the most common being the N-protein or the S-protein.

In a recent population-based study from Spain, different seroprevalences were estimated by a chemiluminescent microparticle immunoassay (CMIA) for detection of anti-N IgG, and a lateral flow immunochromatographic assay (LFIA) for detection of anti-S1 IgG.^3^ Whether the difference in estimated seroprevalence could be explained by dynamic appearance of antibodies targeting the different viral proteins or whether the rapid point-of-care (POC) test did not have as good performance as the CMIA-test, is unknown.

Available serological methods to date either rely on quantitative laboratory-based assays or on qualitative LFIAs. While the lateral flow tests have the advantage of being a POC-analysis where results can be given directly to the patient within minutes from sample collection, rapid POC-tests have been attributed a potential risk of production errors that may result in unreliable performance of the test. There is, however, to our knowledge no study published to date investigating whether the sensitivity and/or specificity varies between production lots of a rapid test. In this study, we therefore aimed to evaluate two separate production lots of one commercially available rapid POC-test (the COVID-19 IgG/IgM Rapid Test Cassette [Zhejiang Orient Gene Biotech Co Ltd, Huzhou, Zhejiang, China]) and its accuracy in identifying anti-S1 IgM/IgG. As reference standard, presence of SARS-CoV-2 S1-specific IgM and IgG in positive controls were verified with an in-house Luminex-based COVID-19 assay (Magpix technology, Luminex Corporation).

## Material and methods

### Serum samples

Positive controls constituted of serum from Swedish COVID-19 patients or convalescents, confirmed positive for SARS-CoV-2 by reverse transcriptase polymerase chain reaction (RT-PCR) and/or serology for SARS-CoV-2 specific IgM and IgG, between March to July 2020. Negative controls constituted of serum from babies (6-12 months old) and randomly selected blood donor sera from the Uppsala Academic Hospital from individuals, without any known history of SARS-CoV-2 infection/COVID-19 and before the COVID-19 pandemic (i.e. collected 2018). Clinical samples that had been deposited in Uppsala Biobank were anonymized and used in accordance with local ethical guidelines. They were all used with informed consent according to the Swedish Biobank law, which allows anonymized diagnostic patient samples to be used for purposes similar to those of the original sampling. The study was approved by the Swedish Ethical Review Authority (2020–02047).

### Reference method

Samples were pre-specified when evaluating the index test. The reference method used was a Luminex-based (Magpix technology, Luminex Corporation) SARS-CoV-2-specific assay, developed in-house and used due to high specificity and sensitivity in detecting SARS-CoV-2 S1-specific IgM and IgG (manuscript in preparation). Samples with confirmed SARS-CoV-2 infection that tested positive for SARS-CoV-2 S1-specific IgG with a median fluorescence intensity (MFI) ≥900 were used as reference for positive IgG-samples. Of those, samples positive for SARS-CoV-2 S1-specific IgM with an MFI ≥700 served as reference for positive IgM-samples. Samples with an MFI <300 were considered negative for anti-S1 IgM and IgG. All samples were from unique individuals and analysed anonymously.

### Index test

The index test (rapid POC-test) used was run according to the manufacturer’s instructions (COVID-19 IgG/IgM Rapid Test Cassette; Product/Model: GCCOV-402a, Lot no. 2003287 [Lot A] and Lot no. 2004156 [Lot B]; Zhejiang Orient Gene Biotech Co Ltd, Huzhou, Zhejiang, China / Healgen Scientific LLC, Houston, USA). This test targets anti-S1 IgM and IgG. Briefly, 5 μl of serum were added to the test slide, followed by 80 μl of the buffer provided in the kit. The results were read after 10 minutes (max 15 min) by the naked eye. Test results were blinded to the assessors of the index test, as well as to the assessors of the reference standard.

### Analysis

Only index tests in which the control line changed colour were regarded as valid (no test was excluded from analysis). If a line was observed for IgM and/or IgG, the test was considered positive. The intensity of the colour was not judged. Sample sizes were determined in order to comply with the recommendations from the Swedish Public Health Agency for validation of COVID-19 related serological assays. The sensitivity of the index test was calculated as the proportion of index positives among reference positives, and specificity as the proportion of index negatives among reference negatives. The Wilson Score method was used to calculate 95% confidence intervals (95% CI). Analyses were performed with STATA v13.1 (StataCorp, Texas, USA). Plots for positive and negative predictive value (PPV; NPV) as a function of prevalence were created in R v4.0.0 (R Core Team, 2020). Reporting of results are made according to the 2015 Standards for Reporting Diagnostic Accuracy (STARD) statement.^8^

## Results

### Selected serum samples

One hundred serum samples, confirmed by RT-PCR (n=90) and/or by serology (n=89), had an MFI ≥900 (range: 998 – 6,477) for anti-S1 IgG. Of those, 74 had an MFI ≥700 (range: 738 – 5,916) for anti-S1 IgM. None of the 200 samples from 2018 gathered from the biobank tested positive for anti-S1 IgM and IgG (MFI <300). A flow diagram of sampling is depicted in Figure 1.

**Figure 1.**
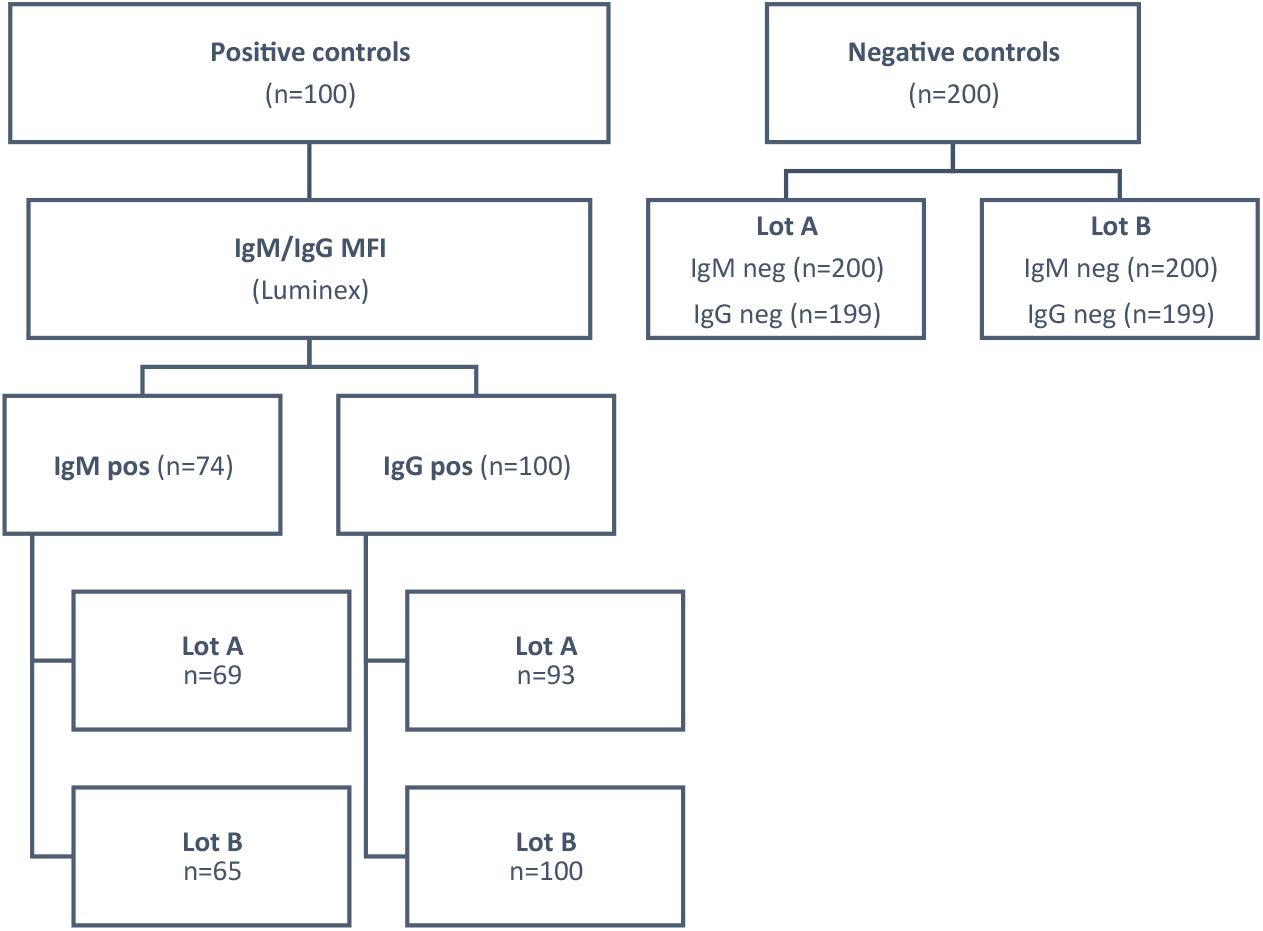
Flow diagram of sampling.

### Index test

With the index test, none of the 200 negative sera from blood donors and babies tested IgM positive (0/200, 0% [95% CI: 0.0-1.9%]), while one tested IgG positive (1/200, 0.5 %, [95% CI: 0.1-2.8%]). This was the case in both production lots tested (Tables 1 and 2). The single IgG-positive sample was re-analysed and remained IgG positive in the second test in both lots. Of the 74 IgM-positive samples, five tested IgM negative (6.8% [95% CI: 2.9-14.9%]) in Lot A and nine (12.2% [95% CI: 6.5-21.5%]) in Lot B. Of the 100 IgG-positive samples, seven tested IgG negative (7.0% [95% CI: 3.4-13.7%]) in Lot A while none tested IgG negative in Lot B (0% [95% CI: 0.0% to 3.7%]). The MFI of the false IgG negatives in Lot A ranged between 998 – 3,453. The MFI of the false IgM negatives ranged 761 – 1,340 and 738 – 1,641 for Lot A and B, respectively.

**Table 1.** Results for serum samples with SARS-CoV-2 specific anti-receptor binding domain of spike protein IgM, confirmed with Luminex technology, and pre-COVID-19 negative controls for two production lots of a rapid point-of-care test.

| Reference method | Index test |  |  |  |  |  |
| --- | --- | --- | --- | --- | --- | --- |
|  | Lot A |  |  | Lot B |  |  |
|  | IgM*<br>Positive | IgM<br>Negative | Total | IgM<br>Positive | IgM<br>Negative | Total |
| **IgM Positive** | 69 | 5 | 74 | 65 | 9 | 74 |
| **IgM Negative** | 0 | 200 | 200 | 0 | 200 | 200 |
| **Total** | 69 | 205 | 274 | 65 | 209 | 274 |

\*Immunoglobulin M

**Table 2.** Results for serum samples with SARS-CoV-2 specific anti-receptor binding domain of spike protein IgG, confirmed by Luminex technology, and pre-COVID-19 negative controls for two production lots of a rapid point-of-care test.

| Reference method | Index test |  |  |  |  |  |
| --- | --- | --- | --- | --- | --- | --- |
|  | Lot A |  |  | Lot B |  |  |
|  | IgG*<br>Positive | IgG<br>Negative | Total | IgG<br>Positive | IgG<br>Negative | Total |
| **IgG Positive** | 93 | 7 | 100 | 100 | 0 | 100 |
| **IgG Negative** | 1 | 199 | 200 | 1 | 199 | 200 |
| **Total** | 94 | 206 | 300 | 101 | 199 | 300 |

\*Immunoglobulin G

### Sensitivity, specificity and predictive values

Based on the results described above and summarized in Tables 1 and 2, the index test had a sensitivity of 87.8% and 93.2% for IgM and 93.0% and 100.0% for IgG (Table 3). The test exhibited an overall specificity of 100.0% for IgM and 99.5% for IgG. The PPV (probability of have been infected and having antibodies given a positive test result) was 100% for IgM and 98.9% and 99.0% for IgG. The NPV (probability of not yet been infected and not having antibodies given a negative test result) was 95.7% and 97.6% for IgM, and 96.6% and 100.0% for IgG. For both antibody types, PPV and NPV remained high over a broad range of prevalence (Figure 2).

**Table 3.** Performance of two production lots of a rapid IgM/IgG test cassette, evaluated using reference samples confirmed by Luminex technology.

| Index test performance |  |  |  |  |
| --- | --- | --- | --- | --- |
|  | IgM |  | IgG |  |
|  | n | % (95% CI) | n | % (95% CI) |
| **Lot A** |  |  |  |  |
| Sensitivity | 69/74 | 93.2 (85.1 – 97.1) | 93/100 | 93.0 (86.3 – 96.6) |
| Specificity | 200/200 | 100.0 (98.1 – 100.0) | 199/200 | 99.5 (97.2 – 99.9) |
| PPV | 69/69 | 100.0 (94.7 – 100.0) | 93/94 | 98.9 (94.2 – 99.8) |
| NPV | 200/205 | 97.6 (94.4 – 99.0) | 199/206 | 96.6 (93.2 – 98.3) |
| **Lot B** |  |  |  |  |
| Sensitivity | 65/74 | 87.8 (78.5 – 93.5) | 100/100 | 100.0 (96.3 – 100.0) |
| Specificity | 200/200 | 100.0 (98.1 – 100.0) | 199/200 | 99.5 (97.2 – 99.9) |
| PPV | 65/65 | 100.0 (94.4 – 100.0) | 100/101 | 99.0 (94.6 – 99.8) |
| NPV | 200/209 | 95.7 (92.0 – 97.7) | 199/199 | 100.0 (98.1 – 100.0) |

The Wilson Score method was used to calculate confidence intervals for proportions. CI indicates confidence interval; IgG, immunoglobulin G; IgM, immunoglobulin M; PPV, positive predictive value; NPV, negative predictive value.

**Figure 2.**
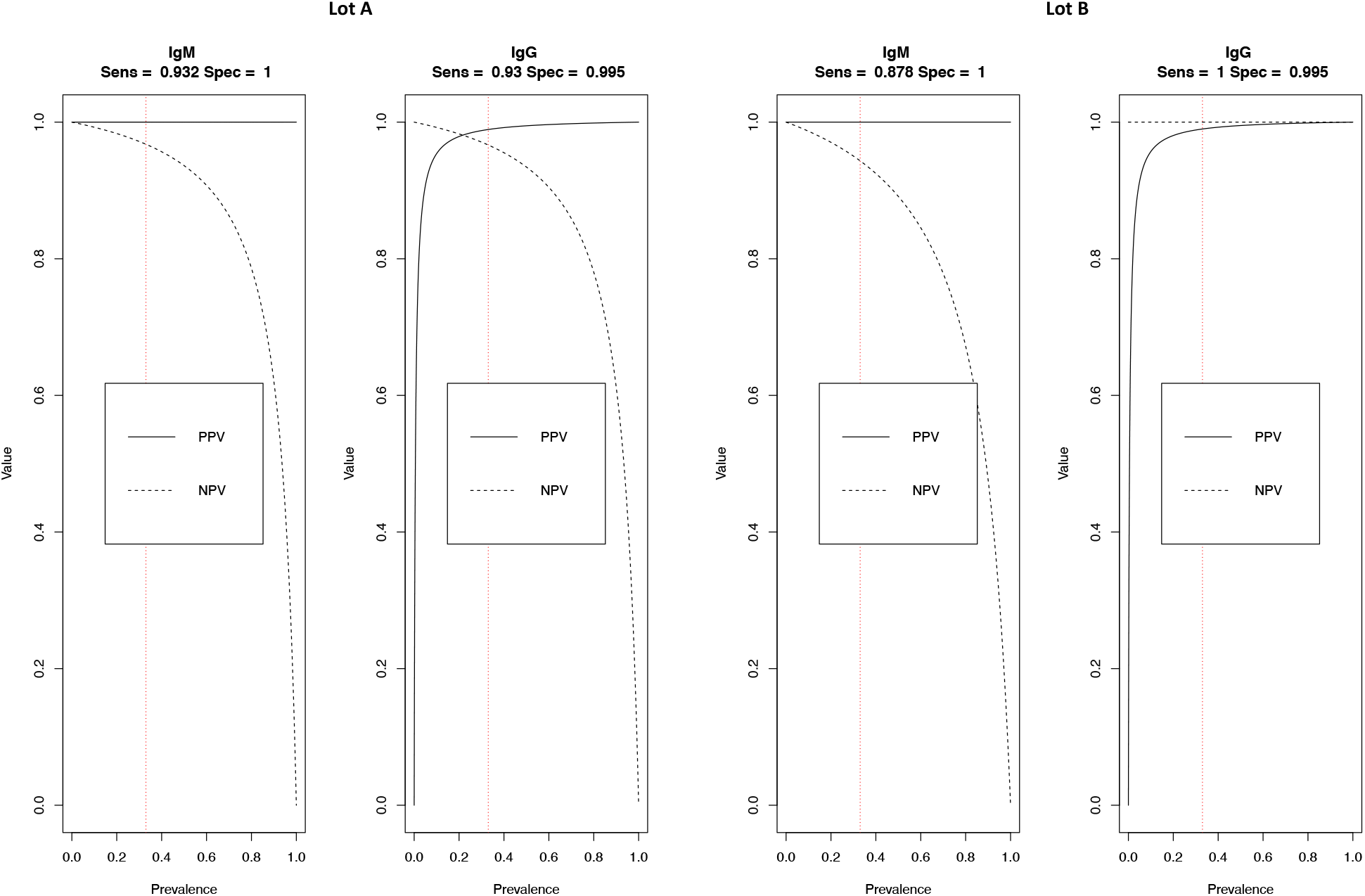
Positive and negative predictive values (PPV; NPV) for two production lots of a rapid point-of-care test as a function of prevalence. Dotted red line indicates study prevalence (33%; 100/300).

## Discussion

In this study, we evaluated different production lots of a commercially available rapid POC-test for detection of IgM and IgG against the S1-protein of SARS-CoV-2. The test displayed a high sensitivity and specificity when compared against a Luminex-based (Magpix technology) SARS-CoV-2-specific assay. Comparing two different production lots, the sensitivity was 87.8% and 93.2% for IgM and 93.0% and 100.0% for IgG. Regarding the specificity, it remained at 100.0% and 99.5% for IgM and IgG, respectively. A high performance of the rapid test has also been reported elsewhere, with a specificity ranging from 97.5% to 100% for IgG and 100% for IgM, and a sensitivity ranging from 96.7% to 98% for IgG and 68% to 100% for IgM.^10^ The Swedish Public Health Agency has warranted a IgG specificity of ≥99.5% and a sensitivity of ≥90% for COVID-19 related serological assays to be recommended as an assay for assessment of antibody presence on an individual level.^11^ In this study, both investigated production lots of the investigated POC-test reached that standard and may, thus, be suitable for antibody screening in Sweden.

Differences in sensitivity of the two production lots in the current study might be explained by small differences in the amount of antigen added during production of the rapid test, which could lead to a weaker qualitative response and, thus, effecting the sensitivity of the test. Different amounts, and not varying quality, of the antigen would also explain why the test was still highly specific in both lots. Previously, we have reported a sensitivity of 69% (20/29) and 93% (27/29) for IgM and IgG respectively for the same rapid POC-test.^12^ In that study, positive controls constituted of serum from PCR-confirmed COVID-19 patients or convalescents with unknown serological response.^12^ Thus, some of the samples serving as positive controls could have been collected in the seronegative window or lost the antibodies, explaining the higher sensitivity observed in the current study.

In a recent population-based study from Spain; more than 51,000 individuals were tested by a CMIA for detection of anti-N IgG and the same rapid POC-test as investigated in the current study, which targets anti-S1 IgM/IgG.^3^ Comparing the two tests, the authors observed different seroprevalences of SARS-CoV-2 specific IgG. Given the high performance in detection of anti-S1 IgM and IgG observed in the current study, a possible explanation for the discrepancy could be that the median seroconversion time, and the antibody peak time, have been observed to occur later for anti-S1 IgM/IgG, as compared to anti-N IgM/IgG.^13,14^ Moreover, anti-S1 IgG levels have been observed to be four times higher during convalescence than in the acute phase in about 40% of patients,^14^ while measurement of only anti-N IgM/IgG have been observed to substantially underestimate the proportion of SARS-CoV-2 infections in general.^15^ Thus, an inter-individual heterogeneity in antigens to which different patients develop antibodies against, or the heterogenous dynamics of antibody appearance, could be a possible explanation to the cumbersome process of inventing a single test that can identify past exposure to the virus with as close to 100% accuracy as possible, despite the worlds weighted efforts. As the current study only aimed to evaluate the performance of the rapid POC-test in detecting anti-S1 IgM/IgG, the non-population-based and predefined study design inherits a weakness in the sense that the POC-test investigated does not detect individuals with antibodies against the N- or other parts of the S-protein. Due to the inter-individual differences in dynamics of these antibody subtypes, it has been suggested that a combination of antibody tests, targeting different viral proteins, may be the best strategy in order to increase sensitivity and/or specificity when screening for SARS-CoV-2 specific antibodies.^3^ However, as anti-S antibodies can block the ACE2-receptor and thus prevent SARS-CoV-2 from infecting the human host,^13^ a positive result from the studied rapid POC-test could indicate immunity against re-infection as long as adequate titers are upheld.

To the best of our knowledge, this is the first study to evaluate the impact of production lots on the performance of a rapid POC-test intended for detection of SARS-CoV-2 specific anti-S1 IgM and IgG. The world is looking for serological assays that can help to decide on relaxation of containment measures and POC-tests are optimal for such purpose given the lower associated costs, easier implementation, and the potentially increased uptake as compared with laboratory-based assays. The fact that the PPV and NPV for IgG remained high over a broad range of prevalence indicates that the investigated rapid POC-test is suitable for screening of SARS-CoV-2 specific IgG antibodies in regions with varying prevalence and during different stages of the pandemic, and therefore could aid in determining containment measures. The observed differences in sensitivity between the two production lots highlight the need for external validation of each production lot before a rapid POC-test is made available on the market.

## Data Availability

All data referred to in the manuscript is available.

## Acknowledgements

This work was supported by the Swedish Research Council (VR, 2017-05807 and 2018-02569). The rapid tests that have enabled this study were donated by the Swedish company Noviral AB (organization number: 559175-7942).

## Disclosure statement

The authors declare no conflicts of interest.

## Notes

### Competing Interest Statement

The authors have declared no competing interest.

### Author Declarations

The study was approved by the Swedish Ethical Review Authority (2020 02047).

